# A Drug Repurposing Strategy for a New Cause of Endometrial Infertility: Unveiling Promising New Treatments

**DOI:** 10.1101/2025.11.04.25339037

**Authors:** Antonio Parraga-Leo, Patricia Sebastian-Leon, Josefa María Sanchez-Reyes, Francisco Jose Sanz, Maria Del Carmen Vidal, Immaculada Sanchez Ribas, Jose Remohi, Antonio Pellicer, Marina Sirota, Patricia Diaz-Gimeno

## Abstract

**STUDY QUESTION:** Which mechanisms of action and candidate drugs can be used to treat endometrial failure caused by molecular alterations rather than endometrial timing?

**SUMMARY ANSWER:** Genistein, pioglitazone, alprostadil, flunisolide, and tenoxicam emerged as potential therapies to treat two molecular causes of endometrial failure not originating in endometrial timing.

**WHAT IS KNOWN ALREADY:** Several studies have described molecular profiles of endometrial failure unrelated to endometrial timing and proposed diagnostic tools based on clinical and transcriptomic data, but effective therapeutic options remain lacking. Hence, there is a pressing need for tailored treatments to enable personalised medicine in endometrial-factor infertility.

**STUDY DESIGN, SIZE, DURATION:** This multicentre prospective study, conducted at 5 fertility clinics in Spain between January 2019 and August 2022, included 192 patients undergoing in vitro fertilisation with hormone replacement therapy, whose endometrial biopsies were collected during the mid-secretory phase.

**PARTICIPANTS/MATERIALS, SETTING, METHODS:** Of 291 endometrial biopsies, 192 met the quality criteria and 161 were classified according to clinical and transcriptomic data using a semi-supervised learning model for prognosis. Before classification, transcriptomic variation related to endometrial timing was corrected using our validated transcriptomic endometrial-dating model. Profiles were analysed using systems pharmacology approaches combining network analysis and reversal signature matching to identify therapeutic drugs capable of reversing molecular disruption. Candidate drugs were grouped by mechanism of action and prioritised by side-effect profile. Selected drugs were validated in endometrial cells through RT-PCR, F-actin staining, and enzyme-linked immunosorbent assays.

**MAIN RESULTS AND THE ROLE OF CHANCE:** Four transcriptomic profiles were identified using artificial intelligence models, each with distinct clinical implications. Two profiles were associated with poor prognosis: clinical miscarriage-associated (CMA, *n* = 27) and biochemical miscarriage-associated (BMA, *n* = 16). CMA was characterised by upregulation of differentiation-related genes and BMA by upregulation of immune-related genes. The 4 profiles were homogeneous in demographic and embryological parameters (age, BMI, and embryo quality), reinforcing their biological relevance. Approved drugs capable of reversing these disrupted expression patterns were identified. Both BMA and CMA were linked to abnormal decidualisation, while BMA also showed immune dysregulation. Genistein and pioglitazone promoted decidualisation in vitro, whereas alprostadil, flunisolide, and tenoxicam inhibited immune responses in endometrial cell cultures, supporting their potential therapeutic role in endometrial failure not originating in endometrial timing.

**LIMITATIONS, REASONS FOR CAUTION:** Although our artificial intelligence-based stratification model revealed clinical and functional differences among profiles, it was designed for drug repurposing rather than predictive diagnosis. A larger, specifically designed study would be required to validate predictive performance and generalisability. Further clinical trials are needed to evaluate the proposed drugs as personalised treatments for this condition.

**WIDER IMPLICATIONS OF THE FINDINGS:** This is the first application of a systems-based drug repurposing strategy in IVF to develop tailored therapeutic interventions. We propose genistein, pioglitazone, alprostadil, flunisolide, and tenoxicam as approved, safe drugs identified through an evidence-based approach that could prevent loss of good-quality embryos and miscarriage due to maternal endometrial factors. These findings, supported by functional in vitro validation, pave the way for future clinical trials advancing personalised medicine in endometrial-factor infertility.

**TRIAL REGISTRATION NUMBER:** Not applicable

## Introduction

The endometrium is a critical tissue in human reproduction. It represents the maternal factor through which the embryo establishes a connection with the uterine surface, leading to placental development and the creation of a suitable environment for foetal growth (Achache and Revel, 2006). This complex process takes place within a narrow period in the menstrual cycle known as the window of implantation (WOI), during which the endometrium is receptive and prepared to accept an embryo (Navot et al., 1991; Wilcox et al., 1999). Following successful implantation, endometrial stromal cells undergo morphological and biochemical changes through a process called decidualisation (Gellersen and Brosens, 2003), which is essential not only for protecting the embryo from maternal immune rejection but also for supplying nutrients required for early embryonic development (Okada et al., 2018). Therefore, the status of the maternal endometrium is essential for reproductive success, and developing strategies focused on endometrial factors is key to improving reproductive outcomes, particularly live birth rates (Strowitzki et al., 2006; Deryabin and Borodkina, 2024).

In recent years, two types of endometrial failure have been identified according to whether it is caused by a problem in endometrial timing, where the endometrium and embryo are not synchronized, or by another cause not related to timing (Sebastian-Leon *et al*., 2018). This second type is known as endometrial failure not originating in endometrial timing, non-timing endometrial failure or independent of endometrial timing, and it refers to a molecular dysfunction of the endometrium during the WOI that results in unsuccessful pregnancy outcomes, including both implantation failures and miscarriages (Sebastian-Leon et al., 2018; Diaz-Gimeno et al., 2022, 2024). Unfortunately, endometrial failure can only be diagnosed clinically based on its symptoms, typically after multiple failed conception attempts—commonly classified as recurrent implantation failure (RIF; Cimadomo et al., 2021) or recurrent pregnancy loss (RPL; European Society of Human Reproduction, 2017). The disparities in clinical classification, along with the heterogeneity and multifactorial nature of this condition, hinder the development of accurate preventive diagnosis and identification of effective treatments (Cimadomo et al., 2021). Nevertheless, integrating high-throughput sequencing with clinical data and artificial intelligence (AI) algorithms offers a promising data-driven approach to better understand and characterise these infertility-related diseases (Díaz-Gimeno et al., 2011, 2017; Koot et al., 2016; Sebastian-Leon et al., 2018; Diaz-Gimeno et al., 2022, 2024).

While advancements in diagnostic tools for dating endometrial biopsies collected in the mid-secretory phase have contributed to a better understanding of endometrial failure (Sebastian-Leon et al., 2018; Devesa-Peiro et al., 2021; Diaz-Gimeno et al., 2022, 2024), the target population and clinical interventions to be applied remain controversial, with no clear clinical benefit demonstrated so far (Simón et al., 2020; Doyle et al., 2021, 2022). Furthermore, the biological basis of endometrial failure not originating in endometrial timing remains poorly understood, despite representing a significant proportion of the population with RIF compared with cases caused by a displaced WOI—highlighting this separate profile as a potentially more relevant cause of poor prognosis (Sebastian-Leon et al., 2018)).

For this reason, in recent years, we have focused our research on non-timing endometrial failure, using two different strategies. One approach involves patient stratification into four profiles using AI models and whole-transcriptomic data, creating an initial classification derived solely from clinical criteria (Sanchez-Reyes et al., 2025). The other leverages a semi-supervised AI framework that integrates both clinical and transcriptomic features across 404 genes to overcome ambiguous clinical endometrial classification, resulting in two different profiles (Diaz-Gimeno et al., 2024). However, despite their potential importance for improving diagnosis of endometrial failure, effective treatments for these molecular alterations remain unknown and are urgently needed. To overcome this gap, the application of systems pharmacology approaches may offer valuable opportunities (Nogales et al., 2022).

Although several computational approaches can be applied to systems pharmacology, transcriptomic-based strategies such as reversal signature matching and network analysis are key components in developing precision medicine approaches (Rodon et al., 2019). These methods leverage gene expression patterns from specific conditions to identify promising targeted treatments (Rodon et al., 2019). One significant application of these methods is drug repurposing, which identifies new indications for already approved drugs and provides notable benefits, including reduced development costs and timelines as well as safer and faster clinical translation (Zhang et al., 2020).

However, maximising the potential of systems pharmacology requires whole-transcriptome analysis to avoid missing key genes that could help us to prioritise mechanisms of action and better stratify the complex, heterogeneous molecular landscape of endometrial failure not caused by WOI displacement (Koot et al., 2016; Sanchez-Reyes et al., 2025). In this study, we aimed to use a whole-transcriptome drug repurposing strategy to identify promising drugs capable of reversing the impaired expression pattern observed in endometrial failure not originating in endometrial timing.

## Materials and methods

### Ethics statement

This study was approved by the Ethics Committee at the Instituto Valenciano de Infertilidad, Valencia, Spain (1706-FIVI-048-PD). Written informed consent was obtained from all recruited participants after full explanation of the study procedures.

### Participants, endometrial biopsy collection, and reproductive outcomes

This multicentre prospective study was conducted between July 2018 and October 2022 at 5 private fertility clinics in Spain. Patients (*N* = 291) were recommended for endometrial evaluation due to medical indications. The patients were undergoing a hormone replacement therapy (HRT) cycle and met the following inclusion criteria: 18– 50 years, with a body mass index (BMI) of 19–30 kg/m2, presenting an endometrial thickness > 6.5 mm with a trilaminar structure in the proliferative phase, and serum levels of oestradiol (E2) > 100 pg/mL and progesterone (P4) < 1 ng/mL on the 10th day of oestrogen treatment. Endometrial biopsies were obtained after an average of 114.47 ± 7.3 h of P4 exposure (range 82–171 h) and pre-processed as previously described (Sanchez-Reyes et al., 2025). The clinical characteristics of the patients were obtained from internal medical records and included age, BMI, reproductive outcomes for each attempt (i.e., pregnancy, live birth, and biochemical or clinical miscarriage), as well as transferred embryo quality (defined in the Supplemental Material).

### Study design

First, endometrial biopsies and clinical characteristics of the population undergoing IVF treatments were collected and recorded. Biopsies meeting quality criteria were sequenced and the transcriptomic variation related to endometrial cyclic tissue changes (the luteal-phase timing effect) was corrected. Subsequently, AI models were used to stratify the population into four profiles according to their clinical and whole-transcriptome patterns, using a semi-supervised strategy (Diaz-Gimeno et al., 2024). The stratified population was then characterised based on its reproductive, transcriptomic, and functional differences. Moreover, transcriptomic differences between profiles were used to prioritise effective and safe treatments through two systems pharmacology methodologies: network analysis and signature matching. Finally, prioritised treatments were validated in vitro in endometrial cell models to confirm their therapeutic potential for poor-prognosis profiles (**Figure 1**).

**Figure 1:**
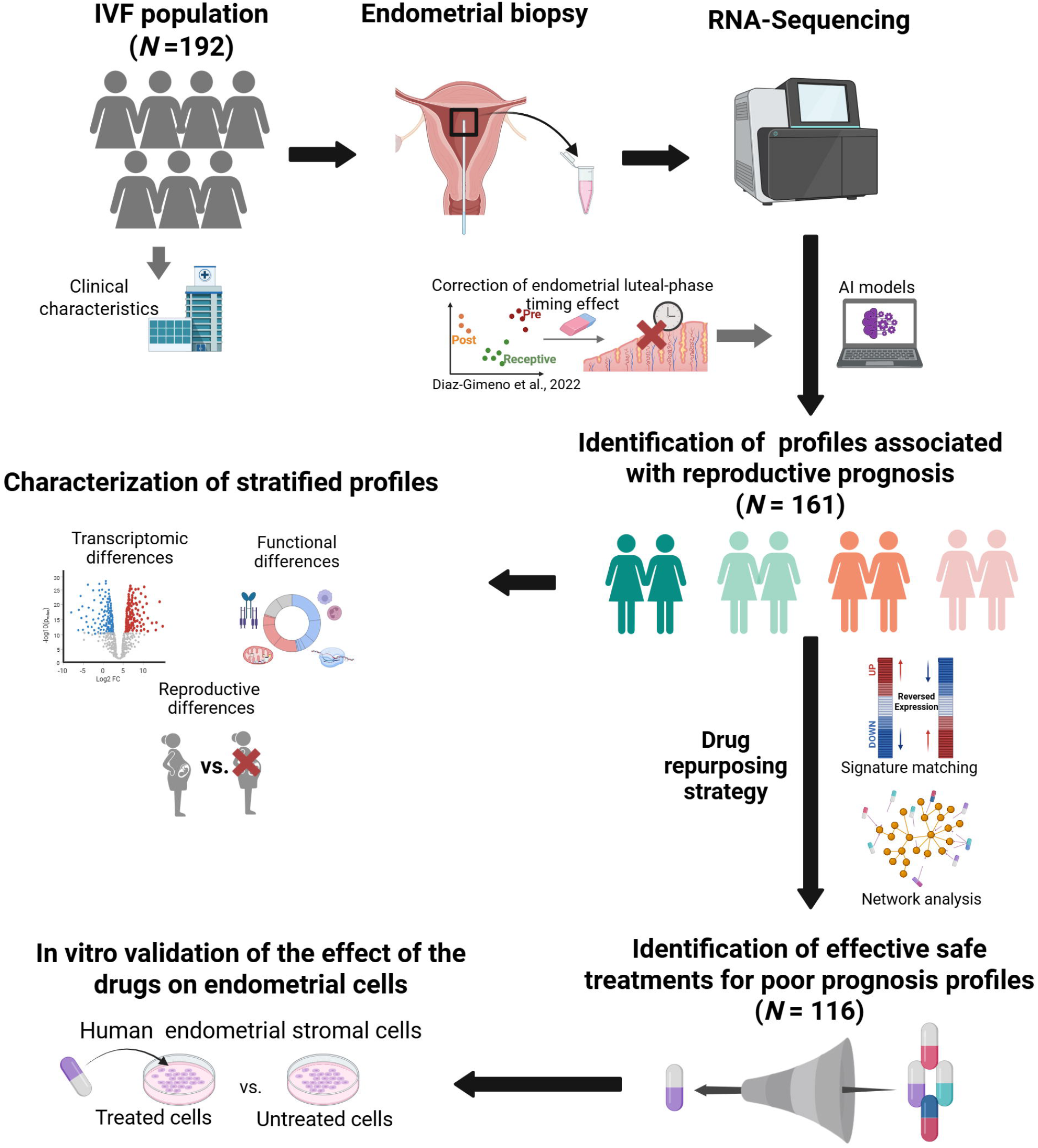
Study design. The diagram provides an overview of the steps performed throughout the study, from patient recruitment to drug validation. First, a population undergoing in vitro fertilisation (IVF) was recruited and relevant clinical data were collected. Endometrial biopsies were then obtained from patients meeting the inclusion criteria and used for gene expression profile analysis. Subsequently, after compensating the endometrial timing effect, an AI model integrating clinical and transcriptomic data was developed to stratify the population into four profiles. These profiles were characterised at the transcriptomic, functional, and reproductive levels. Finally, using gene expression data, potential, already approved drugs were identified for poor-prognosis profiles using two drug repurposing strategies (signature matching and network analysis) and the most promising candidates were assessed in endometrial cell cultures. Abbreviations: IVF, in vitro fertilisation; AI, artificial intelligence.

### Endometrial biopsy gene expression measurement

Coded, anonymised samples were stored in RNAlater (Sigma-Aldrich, Madrid, Spain) at ™80 °C. Total RNA was extracted and samples meeting quality criteria (percentage of RNA fragments larger than 200 nucleotides [DV200] ≥ 70%) were sequenced using an Illumina NextSeq 500/550 system with a paired-end design of 150 cycles and approximately 10 million (M) reads per sample, as previously described (Sanchez-Reyes et al., 2025).

Raw data was evaluated using the FastQC tool (version 0.11.9; Andrews et al., 2010). STAR (version 2.7.3; Dobin et al., 2013) was used to map qualified data. Raw counts were obtained using *featureCounts* from the *Subread* package (version 2.0.3; Liao et al., 2014), and low-quality raw counts (Q < 30) and low-read samples (< 2 M) were filtered out. Counts meeting these quality criteria were normalised using the *limma* R package (version 3.46.0; Ritchie et al., 2015). Genes with low expression (< 1.5 counts per million [CPM] in > 95% of samples) were also filtered out.

Batch effects were detected using principal variance components analysis (PVCA) included in the *PVCA* R package (version 1.36.0; Bushel, 2022) and corrected using linear models implemented in *limma*. Finally, the endometrial-luteal phase timing effect was detected and corrected using our transcriptomic endometrial dating (TED) model (Diaz-Gimeno et al., 2022) as previously described (Devesa-Peiro et al., 2021). Further methodological details are provided in the Supplemental Material.

### Identification of different profiles not originating in endometrial timing associated with reproductive prognosis

Different patient profiles were identified using a two-step stratification employing AI models. Patients were first stratified into two profiles associated with good or poor reproductive prognosis using a semi-supervised AI learning algorithm based on self-labelled techniques, as previously described (Triguero et al., 2015; Diaz-Gimeno et al., 2024). Briefly, this method applies an initial acute clinical classification based on the history of reproductive outcomes in patients who had high-quality embryo transfers and without any uncorrected benign uterine pathologies. Next, AI classification models were implemented to iteratively stratify patients into two profiles associated with a good or poor prognosis based on their transcriptomic profiles, allowing the identification of poor-prognosis profiles associated with endometrial failure.

This stratification represents a personalised medicine strategy that preserves clinical reproductive differences while enhancing transcriptomic resolution to identify homogenous molecular profiles (Diaz-Gimeno et al., 2024). Once patients were divided into poor- and good-prognosis profiles, a *k*-means clustering (Hartigan and Wong, 1979; *k* = 2) unsupervised learning methodology was used within each profile to identify distinct transcriptomic patterns within each prognosis group. This process resulted in a total of four prognosis profiles: two associated with a good reproductive prognosis and two with a poor prognosis (**Supplemental Figure S1A**). Further methodological details are provided in the Supplemental Material.

### Characterisation of stratified profiles

Reproductive outcome rates (i.e., pregnancy rate [PR], live birth rate [LBR], clinical miscarriage rate [CMR], and biochemical miscarriage rate [BMR], as defined in the Supplemental Material) were compared between profiles to characterise their reproductive outcomes. Additionally, to identify genes driving transcriptomic differences between profiles, a differential expression analysis (DEA) was conducted using the *limma* R package. Finally, to conduct a more detailed characterisation, biological functional alterations between profiles were identified through gene set enrichment analysis (GSEA) using the *ClusterProfiler* R package (version 3.18.1; Yu et al., 2012) based on experimental functional annotations included in the Kyoto Encyclopedia of Genes and Genomes (KEGG; Kanehisa and Goto, 2000) and Gene Ontology (GO; Ashburner et al., 2000) databases. Identified biological functions were manually grouped according to their molecular and functional context to facilitate biological interpretation.

### Identification of effective, safe treatments for poor-prognosis profiles based on drug-repurposing techniques

To identify already approved drugs with potential to treat non-timing endometrial disruption and associated with poor-prognosis profiles, we applied a combination of two complementary drug-repurposing methods based on transcriptomic expression and curated protein–protein interactions (**Supplemental Figure S1B**).

Firstly, drugs with potential therapeutic effects for profiles associated with poor reproductive prognosis were obtained using network analysis. DEGs between poor- and good-prognosis profiles were mapped onto the human interactome (see Supplemental Material for more details) to identify disease–protein modules through the KeyPathwayMiner (Mechteridis et al., 2021). Subsequently, the modules were merged into a unique disease network.

Next, to obtain all the approved drugs with at least one target in the disease network and to rank them based on their closeness-centrality scores, the NeDRex tool was used (Sadegh et al., 2021). Finally, to evaluate the therapeutic-effect potential of the identified drugs, the proximity between each drug and its therapeutic targets in the disease network was assessed to exclude drugs without a significant predicted effect on the evaluated profile (Guney et al., 2016; Supplemental Material).

Secondly, to identify drugs with a potential reverse-expression pattern, signature matching techniques were applied. DEG expression patterns between poor- and good-prognosis profiles were combined with drug-induced expression profiles retrieved from the Connectivity Map (CMap) database (Lamb, 2007) to generate a reversal score indicating the potential of each drug to reverse the expression pattern observed in the poor-prognosis profile (Sirota et al., 2011; see Supplemental Material for more details).

Subsequently, both scores were combined to rank drugs according to their predicted efficacy across both approaches, as described in the Supplemental Material. Finally, to prioritise drugs demonstrated to be safe in pregnant individuals, pharmacological information was retrieved from the DrugBank database (Wishart et al., 2018) to confirm their approval for human use, also annotating any adverse effects (especially teratogenic risks) from DailyMed information (“DailyMed,” 2023).

### In vitro validation of the effects of the identified drugs on endometrial cells

Potential drugs identified through drug-repurposing methodologies were grouped according to their mechanisms of action. Since pharmacological action is tissue-dependent, drugs with the best combined scores within each mechanism of action, expressed in the endometrium—based on our dataset and the Human Protein Atlas (Uhlén et al., 2015), and deemed safe for pregnant individuals were prioritised for in vitro validation in immortalised human endometrial stromal cell (HESC) lines.

Subsequently, the optimal concentration of each selected drug was determined through a cell-viability assay (CellTiter 96^®^ Aqueous One Solution Cell Proliferation Assay, G3582, Promega, USA), following the manufacturer’s instructions. HESCs were seeded in 96-well plates at a density of 1.5×105 cells/well and cultivated for 24 h in Dulbecco’s Modified Eagle Medium/Nutrient Mixture F-12 (DMEM/F12) containing 10% (v/v) foetal bovine serum supplemented with 0.2% fungizone and 0.2% penicillin/streptomycin, with increasing concentrations of the evaluated drug or DMSO (vehicle).

Absorbance was measured with a SpectraMax 190 plate reader (Molecular Devices) at 490 nm. Concentrations yielding > 80% cell viability were considered non-cytotoxic, according to ISO 10993-5 standards (Oh et al., 2021; Barbosa et al., 2022). If multiple concentrations exceeded this threshold, previously reported concentrations were used; otherwise, the highest non-cytotoxic concentration was selected.

Next, endometrial cells were cultured with a decidualisation medium as previously described (Salsano et al., 2017). Prolactin and insulin-like growth factor-binding protein 1 (IGFBP-1) levels were qualified using a Human Prolactin ELISA Kit (NBP2-60128, Biotechne, Ireland) and the Human IGFBP-1 ELISA Kit (KA5610, Tebubio, Spain), respectively, following the manufacturer’s protocols. Cell morphology was then visualised using rhodamine–phalloidin staining (Salsano et al., 2021) using an Axio Vert A1 inverted microscope (ZEISS) equipped with a digital camera.

Finally, an in vitro model of lipopolysaccharide (LPS)-induced inflammation was applied as previously described (Zhao et al., 2019). mRNA levels of pro-inflammatory cytokines and inflammation-related genes—interleukin-6 (*IL-6*), tumor necrosis factor (*TNF*), and cyclooxygenase-2 (*COX-2*)—were quantified using quantitative PCR (qPCR), using the primers listed in **Supplementary Table S1**. RNA was extracted using a miRNeasy Mini Kit (Qiagen, Germany) and reverse-transcribed into cDNA using a PrimeScript reagent kit (TAKARA, Japan), according to the manufacturer’s instructions. Relative mRNA expression levels were calculated using the 2™ΔΔCt method (Livak and Schmittgen, 2001).

### Statistical analysis

Experimental and clinical characteristics of patients in each profile were described using the mean and standard deviation for continuous variables and counts and percentages for categorical variables. For continuous variables, the homogeneity across profiles was evaluated using Wilcoxon rank-sum or Student *t*-tests, depending on data normality, while Fisher exact tests were employed for discrete variables. This approach minimised any potential bias in the results associated with confounding variables that were not the main focus of this study. All graphical results were generated using the *ggplot2* R package (version 3.5.1; Wickham, 2009). *P*-values were adjusted using the false discovery rate (FDR) method, establishing a significance threshold of FDR < 0.05.

## Results

### The clinical implications of transcriptomic profiles associated with endometrial failure not originating in endometrial timing

A total of 291 endometrial samples were collected. Of these, 40 were excluded because no clinical follow-up was possible; 56 because the RNA obtained was low-quality or insufficient; and 3 because they had a low number of sequencing reads (< 4 M). Consequently, 192 samples were considered for downstream analyses. Details about the exploratory analysis and correction of batch effects and endometrial luteal-phase timing are included in the Supplemental Material.

To stratify the cohort, first, an initial binary classification was generated comprising 69 patients categorised as pathological-like (*n* = 26) and fertile-like (*n* = 43), leaving 123 (64.1%) patients from the initial population unclassified. After applying this first step, 161 (83.9%) patients could be classified as having a poor (*n* = 43) or good (*n* = 118) reproductive prognosis, while 31 (16.1%) remained unclassified. The performance of the semi-supervised model showed an accuracy of 0.97, sensitivity of 1.00, and specificity of 0.96. Interestingly, patients with a poor prognosis exhibited significantly lower pregnancy and live birth rates and significantly higher clinical and biochemical miscarriage rates, supporting the clinical validity of both profiles (see Supplemental Material for details).

Next, in the second stratification step, patients with a good or poor prognosis were each divided into two groups, yielding a total of four molecular profiles that were homogeneous for clinical and demographic parameters including age, BMI, patient aetiology, endometrial timing, and embryo quality in the first single embryo transfer after biopsy collection (**Supplemental Table S2**). Notably, one of the poor-prognosis transcriptomic profiles—clinical miscarriage associated (CMA, *n* = 27)—had the highest clinical miscarriage rate (50.0%), while the other—biochemical miscarriage associated (BMA, *n* = 16)—showed the highest biochemical miscarriage rate (66.7%), enabling us to associate them with different miscarriage types (**Figure 2A**). Moreover, one of the good-prognosis profiles, the live birth-associated (LBA, *n* = 74) group, exhibited the highest live birth rate (95.2%), whereas the pregnancy-associated (PA, *n* = 44) group showed the highest pregnancy rate (70.7%), albeit with a reduced the live birth rate compared to the LBA profile (79.3% vs. 95.2%, *p* = 0.0246). This finding indicates higher miscarriage rates in the PA profile compared to the LBA one (**Figure 2A**).

**Figure 2:**
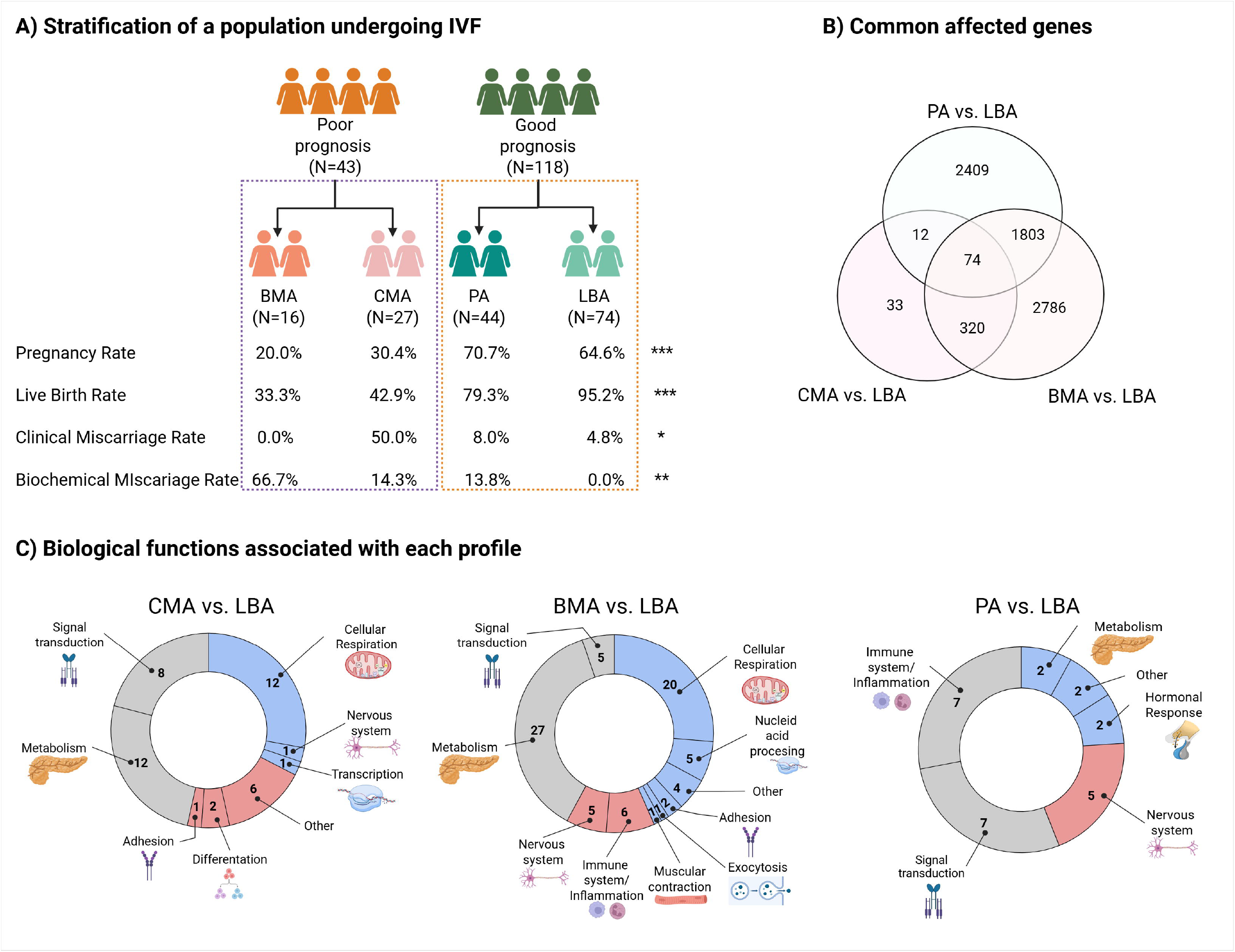
**(A) Stratification of endometrial failure not originating in endometrial timing and associated clinical outcomes**. Four molecular profiles with significant differences in pregnancy, live birth, clinical, and biochemical miscarriage rates were identified. **(B) Common affected genes**. Genes shared across profiles with different clinical symptoms associated with endometrial failure not originating in endometrial timing. The Venn diagram illustrates the differentially expressed genes identified in each comparison. **(C) Biological functions**. Doughnut plots show the biological functions obtained from functional analyses for each molecular profile comparison. The colours indicate whether biological functions were upregulated (red), downregulated (blue), or both (grey) relative to the LBA profile. All the biological functions represented were statistically significant (FDR < 0.05). Abbreviations: BMA, biochemical miscarriage-associated; CMA, clinical miscarriage-associated; PA, pregnancy-associated; LBA, live birth-associated; ***, *p* < 0.001; **, *p* < 0.01; \**p* < 0.05.

Considering the profile with the highest live birth rate (LBA) as the gold standard of the healthiest endometrium, 439 genes were differentially expressed in the profile with the highest clinical miscarriage rate (CMA vs. LBA); 4,983 genes in the profile with the highest biochemical miscarriage rate (BMA vs. LBA); and 4,928 genes with the good-prognosis profile with the lower live birth rate (PA vs. LBA; **Supplemental Figure S2**).

In addition, 74 genes were shared across all the profiles (**Supplemental Table S3**), and 394 shared genes were found between the poor-prognosis profiles, indicating that the CMA profile shared 89.8% of its affected genes with the BMA one. This percentage was lower compared to the PA profile, with only 19.6% (*n* = 86, **Figure 2B**). Moreover, the biological processes underlying each identified profile revealed 44 significantly enriched biological functions, mainly involved in metabolism and signal transduction, in the profile associated with clinical miscarriage (CMA vs. LBA, **Figure 2C**), showing downregulation of cellular respiration and upregulation of differentiation-related pathways.

In total, 76 biological functions were significantly associated with metabolic and signalling alterations, together with heightened inflammatory and immune responses, in the biochemical-pregnancy profile (BMA vs. LBA, **Figure 2C**). Finally, the good-prognosis profile with a lower live birth rate (PA vs. LBA) showed upregulation of nervous system-related pathways along with reduced hormone response (**Figure 2C**). A detailed report of the remaining profile-to-profile comparisons is provided in the Supplemental Material. Similar biological functions were observed in all comparisons using the LBA profile as the reference standard.

### Identification of effective and safe treatments for poor-prognosis profiles

After applying drug repurposing methods, a total of 59 drugs were identified for the CMA profile and 129 for the BMA profile (**Supplemental Table S4**; for details see the **Supplemental Material**). To ensure the safety of the proposed drugs for pregnant individuals, both adverse effects and teratogenic risk were considered (**Supplemental Table S5**), yielding 34 potential drugs for the CMA profile and 82 for the BMA profile (**Supplemental Table S4**, in bold).

Among these, metixene, a muscarinic receptor antagonist used as an antiparkinsonian agent, showed the best score for treating clinical miscarriage. Regarding biochemical miscarriage, tenoxicam, a prostaglandin G/H synthase-2 inhibitor with anti-inflammatory and analgesic properties, achieved the highest score. Interestingly, we also identified several nutraceuticals—including genistein, resveratrol, adenosine phosphate, and calcifediol—which are bioactive compounds commonly used as dietary supplements with health benefits and favourable safety profiles.

To prioritise candidate drugs, we focused on mechanisms of action shared by many different drugs. In particular, for the CMA profile, we selected metixene (an antimuscarinic), zuclopenthixol (a serotonin antagonist), methyldopa (an alpha-adrenergic binder), pioglitazone (a peroxisome proliferator-activated receptor agonist), genistein (an oestrogen receptor binder), terfenadine (an antihistamine), and resveratrol (a nuclear receptor binder), as shown in **Figure 3A** and **Supplemental Table S4**. Topoisomerase inhibitors were excluded because of their known hazardous side effects, while antidopaminergic drugs were not prioritised because their target receptors are not expressed in endometrium according to Human Protein Atlas.

**Figure 3.**
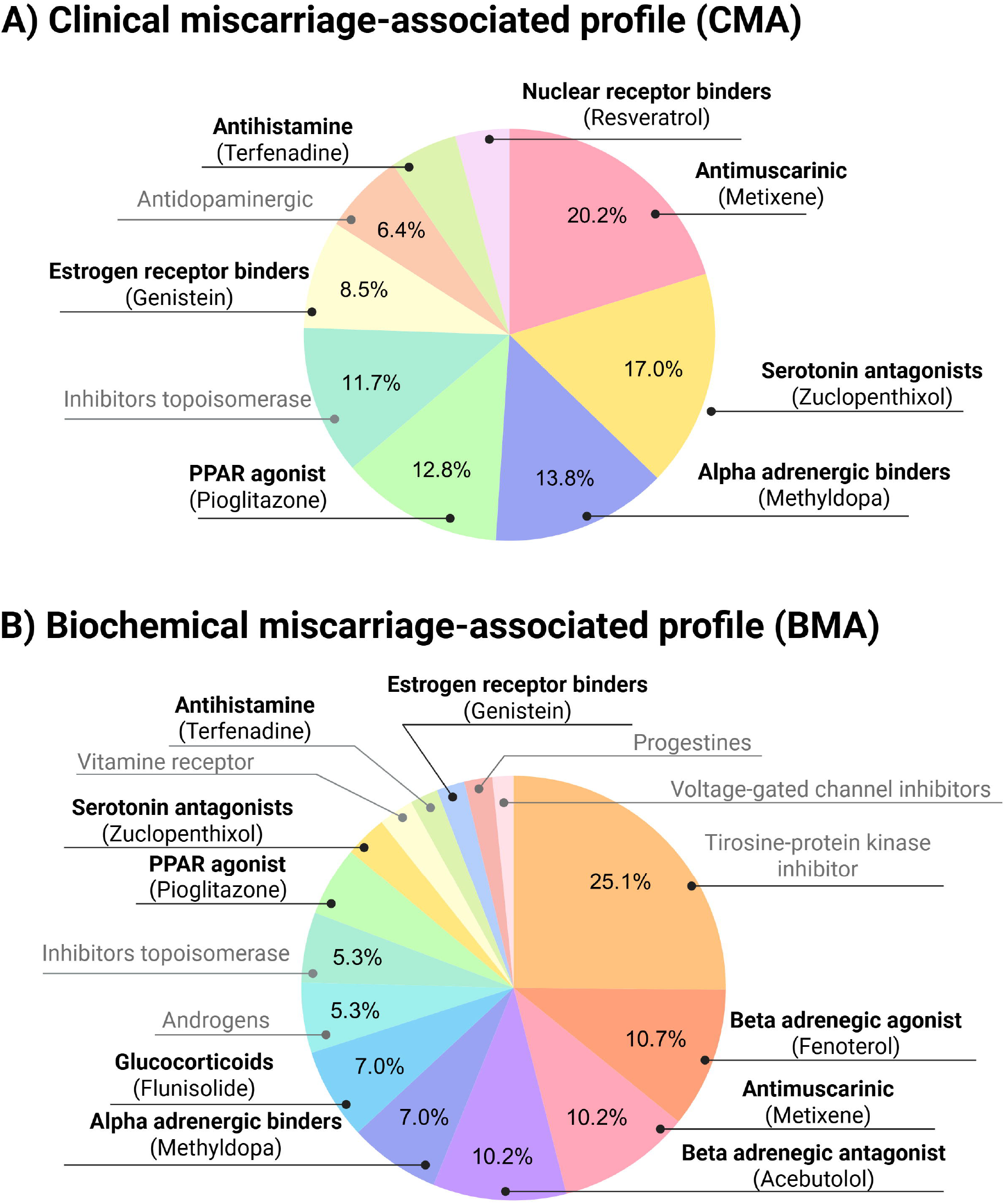
Selected drugs associated with biochemical and clinical miscarriages for in vitro validation. The most frequent mechanisms of action among drugs identified for the CMA **(A)** and BMA **(B)** profiles are shown. The pie charts illustrate the distribution of the main mechanisms of action shared by repurposed drugs predicted to treat each profile. The drug selected for validation, based on its final score, is indicated in parentheses. Mechanisms highlighted in light grey were not selected for further validation. Abbreviations: BMA, biochemical miscarriage-associated; CMA, clinical miscarriage-associated.

Interestingly, biochemical and clinical miscarriage profiles showed notable overlap, except for nuclear-receptor binders. Additionally, the BMA profile presented unique mechanisms of action, including tyrosine-protein kinase inhibitors, beta-adrenergic agonists, beta-adrenergic antagonists, glucocorticoids, androgens, vitamin receptor ligands, progestins, and voltage-gated-channel inhibitors. Nevertheless, we only prioritised drugs associated with mechanisms of action shared with the CMA profile or those with the highest percentage of drugs in the BMA profile, thereby including the majority of mechanisms of action identified in both profiles: fenoterol (a beta-adrenergic agonist), acebutolol (a beta-adrenergic antagonist), and flunisolide (a glucocorticoid) for further validation (**Figure 3B, Supplemental Table S4**).

It is worth noting that although tyrosine-protein kinase inhibition was the mechanism of action with the highest percentage in the BMA profile, none of these drugs were prioritised because of their teratogenic potential. Furthermore, given that functional analysis revealed the immune system as a specifically upregulated process in the BMA profile (**Figure 2C**)—a mechanism frequently linked to biochemical miscarriage in the scientific literature—we also prioritised tenoxicam (a prostaglandin-synthase inhibitor) and alprostadil (a prostaglandin E1 analogue) because of their immunomodulatory and anti-inflammatory potentials. The optimal dose for each selected drug is provided in the Supplemental Material.

### Genistein and pioglitazone lead to decidualisation in endometrial cells

After analysing the signalling pathways for the observed mechanisms of action, we noticed that most of these mechanisms in both profiles (CMA and BMA) acted through three main signalling molecules—cAMP, calcium, and AMP-activated protein kinase (AMPK)—suggesting a potential role for decidualisation in both profiles. Consequently, we evaluated the ability of the selected drugs to promote decidualisation. To assess this effect, we examined three established decidualisation biomarkers: cytoskeletal remodelling, prolactin, and IGFBP-1.

Regarding cytoskeleton remodelling, treatment of endometrial stromal cells with genistein or pioglitazone induced them to form the polygonal morphology typical of decidualised cells (**Figure 4A**). Subsequently, we measured IGFBP-1 secretion, observing a significant increase in this biomarker after treatment with genistein and pioglitazone compared with the negative control treated solely with MPA (**Figure 4B**). Nonetheless, IGFB-1 levels remained lower than those obtained using the gold-standard protocol, indicating that these drugs may support the decidualisation process in impaired stromal cells with reduced cAMP levels but might not independently induce decidualisation. Finally, neither drug led to a significant increase in prolactin secretion, although a slight rise was observed following treatment with pioglitazone (**Figure 4C**).

**Figure 4:**
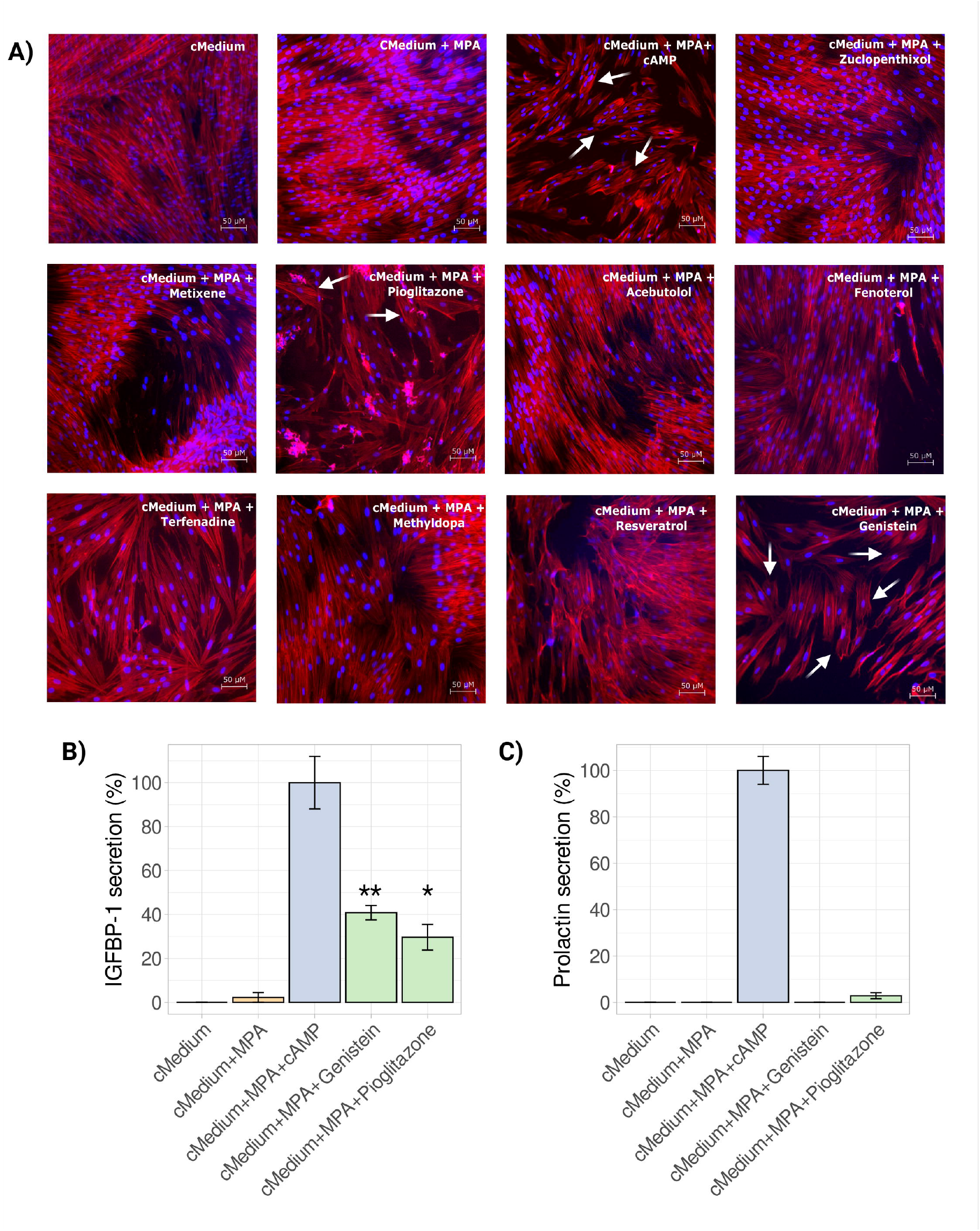
Genistein and Pioglitazone trigger decidualisation in endometrial cells in vitro. Endometrial stromal cells were treated with control medium alone, control medium plus MPA (negative control), control medium plus MPA and cAMP (positive control), or control medium plus MPA and the selected drugs. **(A)** Cytoskeleton remodelling was assessed by observing F-actin filaments stained with phalloidin (red) and nuclei with DAPI (blue). Scale bar: 50 μM. **(B)** and **(C)** IGFBP-1 and prolactin levels were measured by ELISA. Values are expressed as a percentage relative to the positive control (cMedium + MPA + cAMP). Means were compared using the Student’s t-test. Abbreviations: MPA, medroxyprogesterone; cMedium, control medium; cAMP, cyclic AMP. \**p* < 0.05, \*\**p* < 0.01.

### Alprostadil, flunisolide, and tenoxicam reduce the inflammatory response in endometrial tissue

The BMA profile also showed enrichment for drugs related to immune and anti-inflammatory responses; therefore, we evaluated the potential anti-inflammatory effects of alprostadil, flunisolide, and tenoxicam in endometrial cells. After inducing an inflammatory environment in endometrial stromal cell cultures, alprostadil, flunisolide, and tenoxicam all displayed anti-inflammatory effects. Alprostadil did not significantly alter *IL6* gene expression (**Figure 5A**). However, this drug produced a marked and statistically significant decrease in the expression of *COX2* and *TNF* (**Figure 5B, C**).

**Figure 5:**
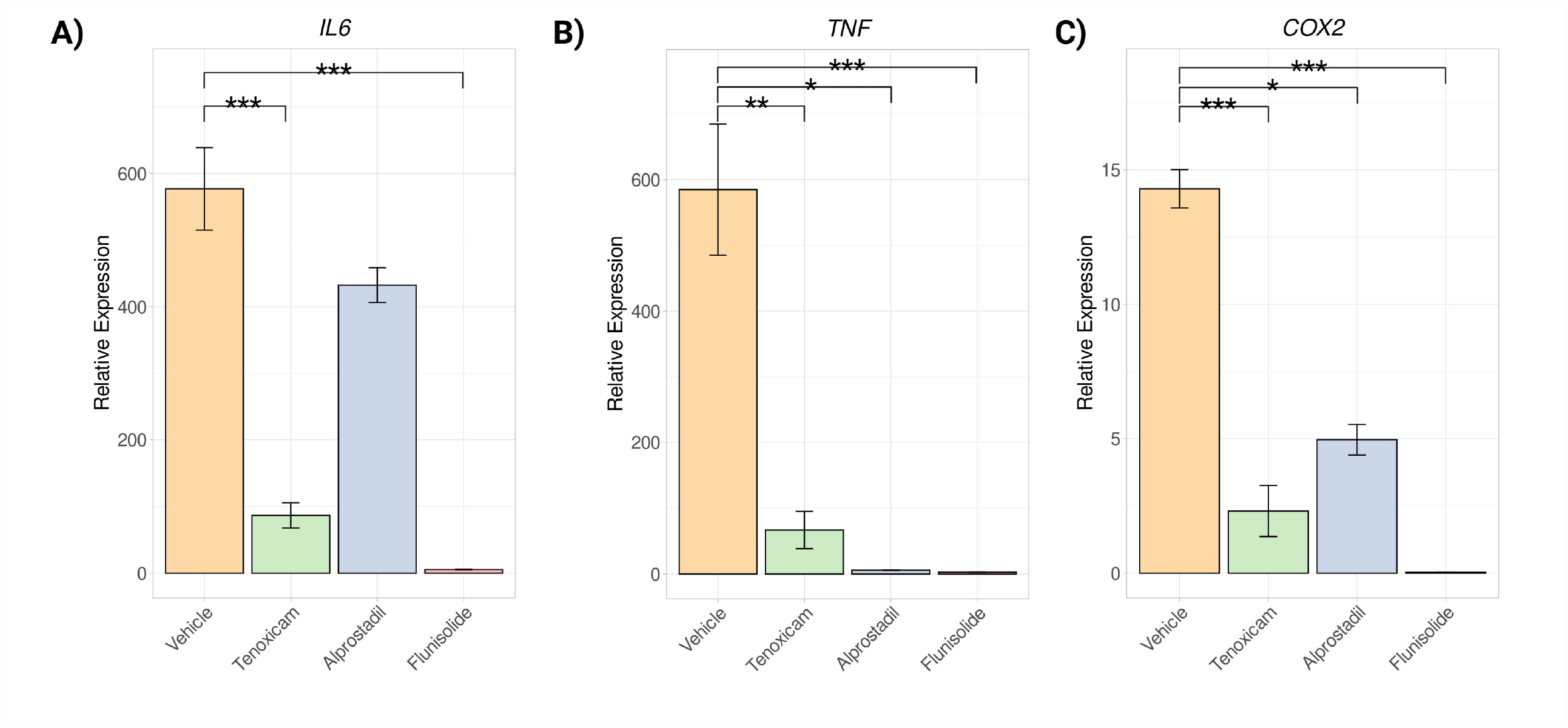
Evaluation of the anti-inflammatory effect of the identified drugs in endometrial stromal cells. Bar plots represent the relative expression levels of three inflammation biomarkers: IL6 **(A)**, *TNF* **(B)**, and *COX2* **(C)**, after treatment with LPS alone (vehicle) or LPS combined with the selected drugs. Abbreviations: Interleukin-6, *IL6*; tumor necrosis factor *TNF*; cyclooxygenase 2, *COX2*. \*\*\**p* < 0.001; **, *p* < 0.01; \**p* < 0.05.

## Discussion

Despite major technological advances and growing knowledge in the field of reproduction, endometrial failure remains a critical concern both in clinical practice and for people trying to conceive. Although several diagnostic tools have been developed, the heterogeneous nature of this condition continues to hinder accurate patient classification and, ultimately, the development of effective therapies. This study aimed to shift the clinical concept of RIF from a symptom-based approach towards caused-based medicine by stratifying the disease into homogeneous molecular profiles using clinical and whole-transcriptome data—an essential step for identifying potential treatments. For the first time, we identified potential drug candidates for treating profiles associated with clinical and biochemical miscarriages resulting from endometrial failure. These compounds showed promising functional effects in endometrial tissue in vitro and were safe for pregnant individuals.

Although previous studies have investigated endometrial failure not originating in endometrial luteal-phase timing, their initial classification for building the machine-learning models was based exclusively on clinical parameters (Koot et al., 2016; Sanchez-Reyes et al., 2025) or their models incorporated only a limited set of genes (Diaz-Gimeno et al., 2024), thereby hindering the stratification of a condition with a highly heterogeneous molecular nature and making it difficult to identify tailored treatments.

To overcome this limitation, in the present study we combined semi-supervised and unsupervised learning approaches to integrate whole-transcriptome information with detailed clinical data, using the latter as a baseline to improve the traditional clinical patient classification. This strategy enabled a more comprehensive stratification of this heterogeneous condition, avoiding the loss of relevant genes during selection and facilitating the identification of DEGs. Moreover, unlike Diaz-Gimeno et al. (2024), we improved the control of embryo quality by establishing an acute clinical classification based exclusively on high-quality embryos, either obtained from donors or confirmed as euploid through preimplantation genetic testing. This approach minimises embryo-related confounding, ensuring that our findings are primarily associated with the endometrial factor.

As expected, and consistent with previous studies based exclusively on clinical data and supervised learning (Koot et al., 2016; Sanchez-Reyes et al., 2025), our approach identified a substantially larger set of genes associated with endometrial disruption. This enhanced the detection of molecular differences and provided strong evidence of the success of our method to define homogeneous molecular profiles with clear clinical relevance in the endometrium of patients undergoing IVF. Nonetheless, it is worth noting that more than 4,000 genes were differently expressed compared with the LBA profile; however, the CMA profile showed fewer transcriptomic differences, with a total of 439 genes. This finding suggests that endometrium in patients with a CMA profile is more similar to that of individuals with a LBA profile and the high clinical miscarriage rate in this group may not be driven solely by endometrial factors. It could also involve embryo–endometrial interactions or embryo-related factors other than ploidy.

Nevertheless, our findings provide clear evidence that endometrium plays an important role in ongoing pregnancy and live birth, as previously reported (Muter et al., 2025). Moreover, we also observed that the CMA profile shared most of its DEGs with that of the BMA profile (89.8%), which may indicate that both profiles exhibit common impaired biological processes. Therefore, a shared therapeutic approach could potentially be developed to simplify and improve clinical management.

Furthermore, no significant differences in demographic or clinical variables were found between any of the identified profiles, reinforcing the robustness of our stratification results. However, although age differences were not statistically significant among groups (*p* = 0.06), an increasing trend was observed, with older age associated with poorer reproductive outcomes. This trend may be explained by the well-documented impact of age on endometrial function, as reported in several studies (Devesa-Peiro et al., 2022; Loid et al., 2024; Marti-Garcia et al., 2024b; Sebastian-Leon et al., 2025). Notably, poor-prognosis profiles showed mean ages of 40 and 41 years, coinciding with the age threshold at which implantation failure and miscarriage rates begin to rise sharply (Sebastian-Leon et al., 2025). Additionally, endometrial luteal-phase timing was not associated with any of the identified profiles (*p* = 0.93), confirming both adequate correction for endometrial progression effects and the independence of these endometrial profiles from endometrial timing-related causes.

The whole-transcriptome differences associated with clinical outcomes also enabled a detailed characterisation of complex molecular profiles with clear clinical relevance, facilitating the identification of promising repurposed drugs capable of reversing the altered expression patterns observed. Indeed, these innovative strategies, which integrate systems pharmacology and transcriptomics, have recently been applied to other reproductive disorders such as endometriosis (Oskotsky et al., 2024; Sanami et al., 2025) and polycystic ovary syndrome (Wu et al., 2022), yielding promising results and further supporting the validity of the methodology used in this study.

Overall, the identified drugs converged on common signalling pathways involved in decidualisation, including AMPK (Hong et al., 2024), cAMP (Gellersen and Brosens, 2003), and calcium signalling (Kusama et al., 2015; Okada et al., 2018). These findings suggest that impaired decidualisation underlies poor-prognosis profiles, supporting the concept that decidualised cells act as biosensors with a dual roles—receptivity and selectivity. This also indicates that endometrial failure is likely driven by defective selectivity, whereby good-quality embryos implant but are subsequently rejected, leading to in high miscarriage rates (Macklon and Brosens, 2014). In addition, specific pathways related to biochemical miscarriage such as anti-inflammatory cascades involving prostaglandins, cyclooxygenases, and glucocorticoids were observed, reinforcing the role of immune-system dysregulation in this profile (Sargent et al., 1988; Colucci, 2019, Sanchez-Reyes et al., 2025).

Analysis of the drug candidates revealed a wide range of receptor targets that could be categorised into specific drug classes. Notably, oestrogen binders were present in both profiles, highlighting a stronger contribution of oestrogen signalling over progesterone in endometrial failure, as previously reported by other authors (Koot et al., 2016; Parraga-Leo et al., 2023; Marti-Garcia et al., 2024a). Although a large number of drugs were identified and made available to the scientific community through our results, we prioritised only the safest drugs with the highest therapeutic potential. Pioglitazone and genistein showed pro-decidualisation effects, which would benefit the impaired decidualisation observed in both poor-prognosis profiles (CMA and BMA).

In addition, we also validated specific drugs for the BMA profile (alprostadil, tenoxicam, and flunisolide) which, through different mechanisms of action (as an anti-inflammatory prostaglandin analogue, COX-2 inhibition, and glucocorticoid activity, respectively) promoted inhibition of inflammation, counteracting the immune-system overactivation observed in this profile. Consequently, these results highlight safe and promising drug candidates with validated effects in endometrial tissue, representing potential therapies for female infertility associated with endometrial failure.

Despite the fact that both poor-prognosis profiles exhibited impaired decidualisation, their underlying molecular mechanisms may differ substantially. Indeed, immune-system dysregulation has been reported to disrupt the decidualisation process, leading to early pregnancy loss (Liu et al., 2024), which could explain the findings observed for the BMA profile. Furthermore, given the larger number of differentially expressed genes and more extensive functional alterations observed in the BMA group, further investigation is warranted to determine whether a single treatment would be sufficient to address this condition or whether a combination therapy would be more effective.

Finally, an interesting finding reported for first time in this study was the good-prognosis profile PA, which was characterised by a lower hormonal response. This feature may explain the reduced live birth rate for this profile compared with the benchmark LBA profile. However, the main objective of this study was to focus on the profiles associated with poor prognosis, which have a greater impact on clinical outcomes. Future studies should therefore aim to propose therapeutic candidates to improve the live birth rate in this group.

Although the findings of this research are promising, several limitations must also be acknowledged. First, the sample size within each group was limited and a larger cohort will be required to validate the generalisability of the results. Second, drug effects were assessed in vitro using cell culture models, which do not capture the complexity of the full endometrial microenvironment. Third, although the identified drugs are already approved, the optimal route of administration for endometrial targeting remains to be determined to maximise therapeutic efficacy. Nevertheless, despite these limitations, it is worth emphasising that our stratification is supported not only by strong clinical correlations but also by molecular evidence. Given that the proposed drugs are already approved and have favourable safety profiles, early-phase clinical trials (phase I or II) could be initiated, facilitating their translation into clinical practice (Zhang et al., 2020).

To conclude, this work introduces an innovative approach in reproductive medicine, enabling the identification of effective drugs through robust drug-repurposing pipelines based on accurate patient stratification, thereby accelerating clinical translation. This approach promotes the development of preventive medicine strategies aimed at minimising the loss of valuable embryos and improving reproductive outcomes. Ultimately, these findings pave the way for future clinical trials and offer new hope to individuals with non-timing endometrial failure endometrial timing, for whom no effective treatment currently exists.

## Data availability

FASTQ files for bulk RNA-seq and raw count matrix were deposited at the Gene Expression Omnibus (GEO) under accession number GSE283409.

## Author roles

A.P.-L.: investigation, visualisation, formal analysis, software, writing – original draft; S.P.-L.: data curation, formal analysis, visualisation, software, writing – original draft; J.M.S-R; investigation, data curation, resources; F.J.S.: investigation, writing – review & editing; M.C.V.: patient recruitment and sample collection; I.S-R: patient recruitment and sample collection; A.P.: patient recruitment, sample collection, and supervision; M.S.; formal analysis, resources, writing – review & editing; P.D.-G.: conceptualisation, methodology, writing – review & editing, supervision, project administration, funding acquisition. All authors have read and approved the final version of the manuscript.

## Acknowledgements

We thank all IVF participants from the IVI clinics of Spain. We also thank Marina Sirota’s research group, especially Tomiko T. Oskotsky for their assistance with the drug repurposing algorithm and their valuable feedback. In addition, we would like to thank Prof. Jan Baumbach and his research group for their guidance in the use of Nedrex.

## Funding

This study was Supported by **Fundación IVI (1706-FIVI-048-PD)**, Instituto de Salud Carlos III (ISCIII, Spanish Ministry of Science and Innovation) co-funded by the European Regional Development Fund ‘A way to make Europe’ (**PI19/00537** [P.D.-G.]) and Generalitat Valenciana, ‘subvenciones concedidas a grupos de investigación emergentes - GE2024 (CIGE2023)’ (**CIGE/2023/117** [F.J.S.]). P.D.-G. is supported by the ISCIII **Miguel Servet** Programme (**CP20/00118**) and co-funded by the EU. A.P.-L. is supported by the Spanish Ministry of Science, Innovation, and Universities (**FPU18/01777**). P.S.-L. and F.J.S. are funded by the ISCIII **Sara Borrell** Programme (**CD21/00132** and **CD23/00032**, respectively), co-funded by the EU. **Marina Sirota:** March of Dimes Prematurity Research Center at UCSF, NIH: P01HD106414, R01HD105256 1R21HD114953-01. All authors declare no conflict of interest.

## Conflict of interest

All authors declare no conflict of interest.

## Supplemental Figures

**Supplemental Figure S1:**
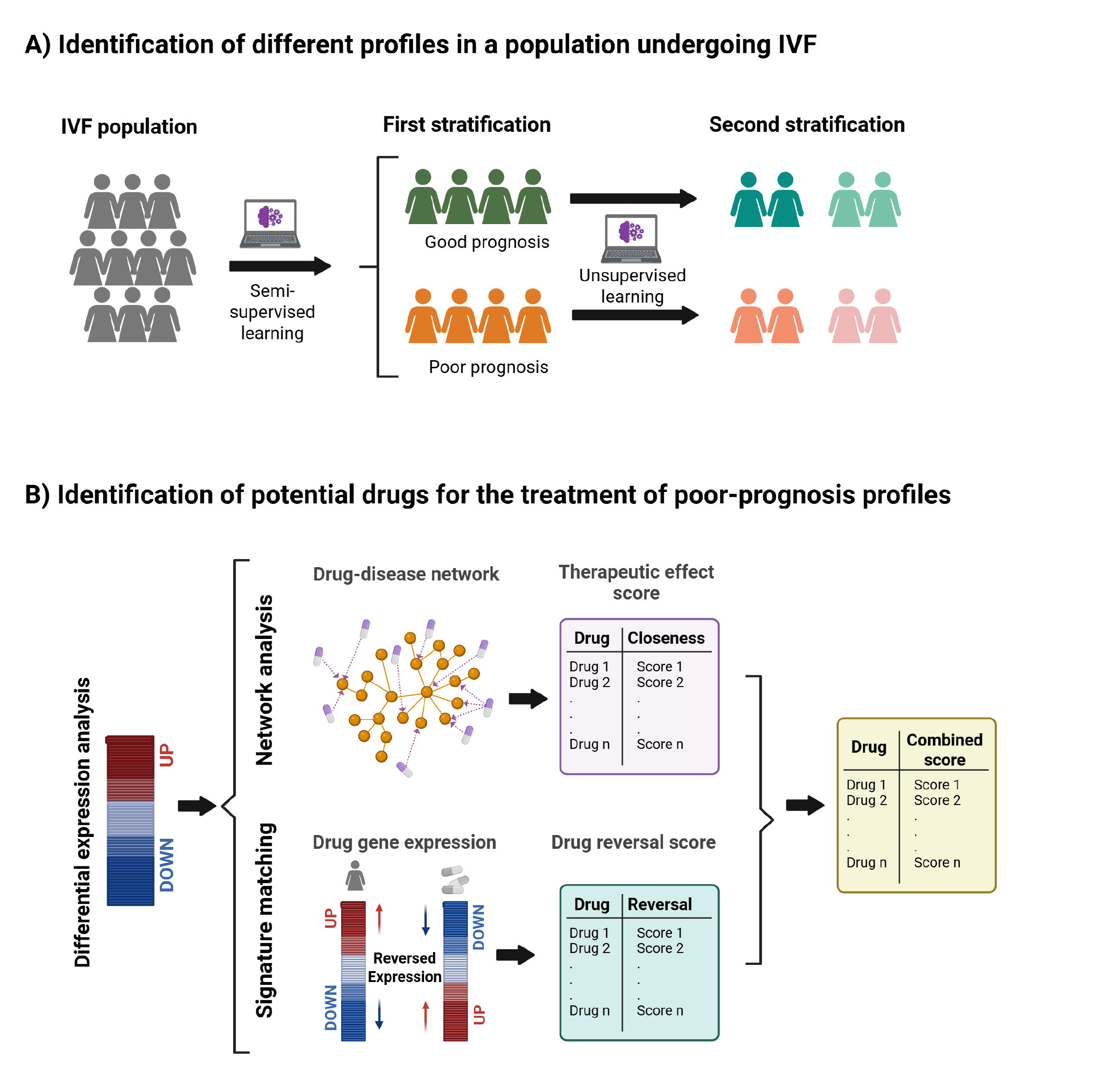
Stratification of endometrial disruption and identification of potential drugs. **(A)** Samples included in the study were first classified into two groups (good- and poor-prognosis profiles) using a semi-supervised learning approach. Afterwards, each identified profile was further stratified through an unsupervised learning method into two additional profiles, yielding four distinct molecular profiles for the studied population. **(B)** Leveraging the gene expression pattern from differentially expressed genes (DEGs), two drug-repurposing strategies were applied. First, a network analysis using the human interactome and information obtained from drug databases was conducted to identify potential drugs and their associated therapeutic-effect scores. Second, a signature-matching approach was applied to identify drugs capable of reversing the impaired gene expression pattern associated with endometrial disruption, yielding a drug-reversal score for each of candidate. Finally, both approaches were merged and the resulting scores were combined.

**Supplemental Figure S2:**
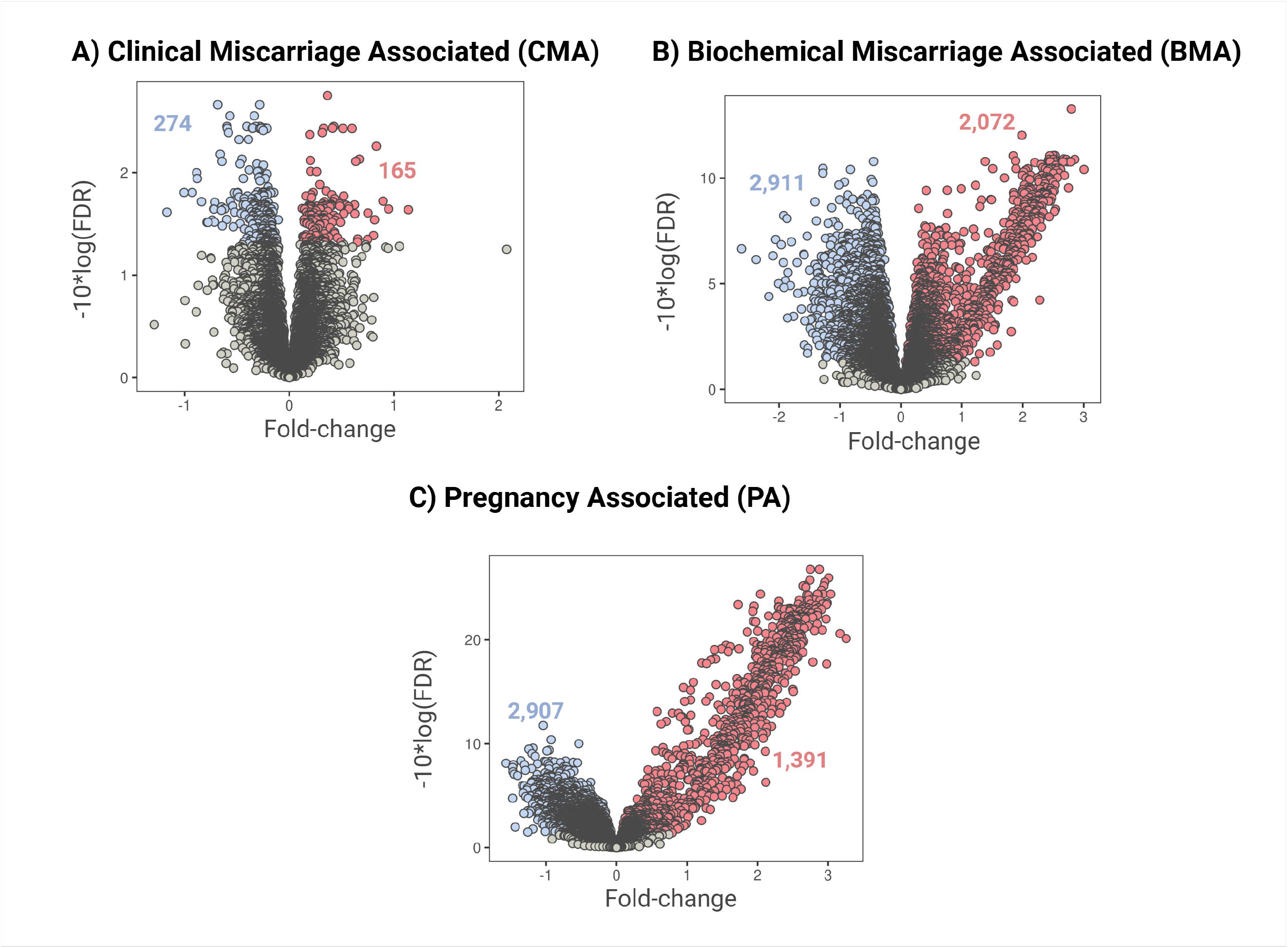
Transcriptomic differences relative to the live birth-associated profile (LBA). **(A)** CMA vs. LBA; **(B)** BMA vs. LBA; **(C)** PA vs. LBA. Volcano plots represent the differential expression analysis results for each comparison. Colours indicate if differentially expressed genes were significantly upregulated (red), significantly downregulated (blue), or non-significant (grey) when compared to the LBA profile. Abbreviations: BMA, biochemical miscarriage-associated; CMA, clinical miscarriage-associated; PA, pregnancy-associated; LBA, live birth-associated; FDR: false discovery rate.

## Supplemental Tables

**Supplementary Table S1:**
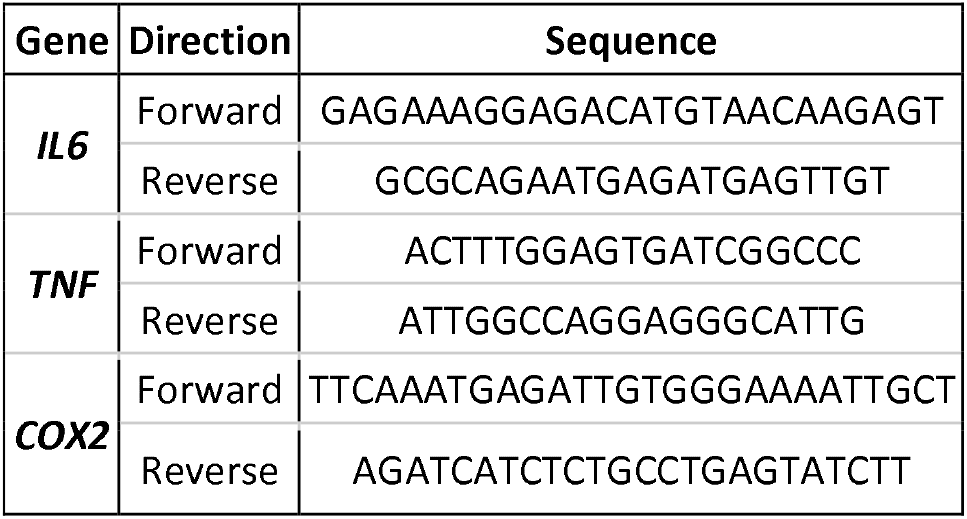
Specific primers used for quantitative reverse-transcription PCR (RT-qPCR). Specific forward and reverse primer sequences employed in the RT-qPCR validation of the interleukin 6 (*IL6*), tumor necrosis factor (*TNF*), and cyclooxygenase-2 (*COX2*) genes.

**Supplementary Table S2.** Summary of the clinical information for each profile. Discrete variables are represented as counts and percentages and continuous variables by their means and standard deviations. Abbreviations: BMA, biochemical miscarriage-associated; CMA, clinical miscarriage-associated; PA, pregnancy-associated; LBA, live birth-associated. N<u>o</u>, number of; BMI, body mass index; BUP, benign uterine pathology; HQE, high-quality embryos; UQE, uncertain-quality embryos; NA, not available.

| Variables |  | CMA | BMA | PA | LBA | *p*-value |
| --- | --- | --- | --- | --- | --- | --- |
| **Nº of patients** |  | 27 | 16 | 44 | 74 | – |
| **Age** |  | 41.56 ± 4.25 | 40.06 ± 5.8 | 39.8 ± 4.88 | 38.62 ± 4.48 | 0.06 |
| **BMI** |  | 23.28 ± 3.81 | 22.59 ± 2.99 | 22.13 ± 3.1 | 23.18 ± 3.33 | 0.32 |
| **Female aetiology** | BUP | 1 (3.70%) | 4 (25.00%) | 10 (22.70%) | 9 (12.20%) | 0.09 |
|  | Normal | 20 (74.10%) | 1 (75.00%) | 30 (68.20%) | 58 (78.40%) |  |
|  | NA | 6 (22.20%) | 0 | 4 (9.10%) | 7 (9.50%) |  |
| **Cycles from biopsy to transfer** |  | 4.13 ± 4.1 | 2.47 ± 1.85 | 3.66 ± 2.86 | 3.85 ± 2.68 | 0.31 |
| **Endometrial luteal-phase timing** | Displaced | 15 (55.6%) | 10 (62.5%) | 23 (52.3%) | 40 (54.1%) | 0.93 |
|  | On time | 12 (44.4%) | 6 (37.5%) | 21 (47.7%) | 34 (45.9%) |  |
| **Embryo quality** | HQE | 23 (85.2%) | 15 (93.8%) | 39 (88.6%) | 63 (85.1%) | 0.94 |
|  | UQE | 4 (14.8%) | 1 (6.2%) | 5 (11.4%) | 9 (12.2%) |  |
|  | NA | 0 (0.0%) | 0 (0.0%) | 0 (0.0%) | 2 (2.7%) |  |

## Notes

### Competing Interest Statement

The authors have declared no competing interest.

### Funding Statement

This study was supported by Fundaci&OACUTEn IVI (1706-FIVI-048-PD), Instituto de Salud Carlos III (ISCIII, Spanish Ministry of Science and Innovation) co-funded by the European Regional Development Fund ‘A way to make Europe’ (PI19/00537 [P.D.-G.]) and Generalitat Valenciana, ‘subvenciones concedidas a grupos de investigac&OACUTEin emergentes - GE2024 (CIGE2023)’ (CIGE/2023/117 [F.J.S.]). P.D.-G. is supported by the ISCIII Miguel Servet Programme (CP20/00118) and co-funded by the EU. A.P.-L. is supported by the Spanish Ministry of Science, Innovation, and Universities (FPU18/01777). P.S.-L. and F.J.S. are funded by the ISCIII Sara Borrell Programme (CD21/00132 and CD23/00032, respectively), co-funded by the EU. Marina Sirota: March of Dimes Prematurity Research Center at UCSF, NIH: P01HD106414, R01HD105256 1R21HD114953-01. All authors declare no conflict of interest.

