## Supplementary Methods for "A Drug Repurposing Strategy for a New Cause of Endometrial Infertility: Unveiling Promising New Treatments"

### Controlling embryo quality for endometrial research for in vitro fertilisation treatments

To evaluate embryo quality, two groups were defined: high-quality embryos (HQEs) and uncertain-quality embryos (UQEs). HQEs were defined as autologous embryos whose euploidy had been confirmed by pre-implantation genetic testing (PGT) or as embryos derived from oocyte donation by women under 35 years of age. UQEs referred to autologous embryos whose euploidy could not be confirmed. We selected only HQEs for the acute clinical classification to ensure optimal model performance. When both HQEs and UQEs were included in subsequent analyses, we verified that the proportion of UQEs did not differ significantly among groups, ensuring that no bias was introduced in assessing the contribution of the endometrial factor to infertility.

### Correction of the effect of endometrial cycle progression

Correction of the effect of endometrial cycle progression was performed to remove transcriptomic variability that could mask key biomarkers associated with endometrial dysfunction. Endometrial variability related with endometrial cycle progression was first identified using the validated Transcriptomic Endometrial Dating (TED) model (Diaz-Gimeno et al., 2022), which classifies samples into four secretory-phase profiles. Based on these classifications, the cycle progression effect was then corrected using linear models implemented in the *limma* R package (Ritchie et al., 2015). This correction effectively removed transcriptomic variability related to endometrial progression while preserving molecular differences associated with endometrial dysfunction—the main objective of this study.

### Semi-supervised learning algorithm for improving clinical diagnosis

This algorithm combines an initial acute clinical classification with artificial intelligence (AI) models to stratify patients into two groups, preserving clinical differences while enhancing molecular discrimination through transcriptomic variation. For the initial acute clinical classification, only patients who had had transfers with high-quality embryos (HQEs) and with no evidence of benign endometrial pathology were considered. The acute clinical classification was performed based on clinical variables, specifically the number of single-embryo transfer attempts and the final reproductive outcome. As a result, the population was divided into pathological-like (PL) patients—those who had never achieved a full-term pregnancy after at least three attempts—and fertile-like (FL) patients, who had achieved a full-term pregnancy after at the first attempt.

Gene-signature selection was subsequently performed using the previously described methodology (Diaz-Gimeno et al., 2022). Briefly, genes were ranked according to the informativity score, calculated with the *CorrelationAttributeEval* algorithm implemented in Weka (version 3.8.2, 2017-12-22; Frank et al., 2017) to distinguish the two groups based on the initial acute classification. To optimise gene selection size, multiple gene sets with increasing signature sizes were generated by adding genes sequentially according to their informativity rank, from highest to lowest. These sets were analysed using Support Vector Machine (SVM; Noble, 2006) and Random Forest (RF; Breiman, 2001) models.

Prediction performance was calculated by using a cross-validation process (default parameters; stratified 5-fold cross-validation, 80:20 split, repeated 100 times) implemented in the Rweka R package (Hornik et al., 2009). The smallest number of genes achieving the highest accuracy in each SVM and RF model was selected, and the larger of the two signatures was considered the final predictive gene-expression signature distinguishing between the acute clinical groups. The probability threshold for accurate classification of a new patient was set at 0.75.

**Evaluation of clinical reproductive outcomes**

The pregnancy rate (PR) was defined as the proportion of positive pregnancy tests obtained after the first transfer following biopsy collection in the patient population undergoing embryo transfer. The live birth rate (LBR) was defined as the number of live births divided by the total number of positive pregnancy tests obtained after the first transfer after biopsy collection. The clinical miscarriage rate (CMR) was defined as the proportion of clinically recognised pregnancy loss before the 20th week of gestation in relation to the total number of pregnancies in which a gestational sac had been visualised. The biochemical pregnancy rate was defined as the proportion of gestational losses after a positive pregnancy test without visualisation of the gestational sac. Differences in proportions between the groups were statistically tested using Fisher exact tests.

**Human interactome**

To build a comprehensive, high-quality human protein–protein interactome (PPI) for downstream network analyses, we conducted a literature-based selection of major databases containing experimentally validated PPIs. Six primary-source databases were initially selected: HIPPIE, BioGRID, HPRD, IntAct, MINT, and HuRI. However, HPRD was excluded due to data unavailability. Raw data were retrieved and formatted in Python (version 3.11), then processed in R to retain only experimentally-validated interactions. All gene and protein identifiers were standardised to Entrez IDs, and discrepancies were manually curated. To remove redundant interactions and ensure consistent data integration, we built individual graphs for each database using the iGraph R package (version 1.2.6; Csardi and Nepusz, 2006) and merged them into a unified interactome. The final PPI served as the foundation for constructing disease-specific molecular networks in subsequent analyses.

**Proximity analysis**

To assess whether the potential therapeutic effectiveness of the identified drugs, a network medicine algorithm known as *Proximity Analysis* was adapted and applied (Guney et al., 2016). This method evaluates the topological proximity between drug targets and disease-associated genes in the human PPI, considering the shortest paths connecting both sets of nodes (drugs and disease genes). To assess statistical significance and ensure that the observed proximity was not due to random chance, a reference distribution was generated by calculating the distances between two randomly selected gene groups 1,000 times. Distances from the disease networks were then standardised to compute a *z*-score and the corresponding *p*-value.

**Reversal score**

For the signature-matching strategy, to avoid the selection of an arbitrary number of genes as input, a workflow using multiple thresholds for adjusted *p*-value and fold change was implemented to create several differentially-expressed gene (DEG) sets. For each comparison, a reversal score was generated using the DEG set and drug-induced gene-expression profiles retrieved from the Connectivity Map database. The reversal score, based on the Kolmogorov–Smirnov statistic, was calculated for each drug–disease signature pair. A permutation analysis was performed to assess significance and only drugs with a reversal score $< 0$ and an adjusted *p*-value $\leq 0.05$ were retained. Because multiple DEG sets were analysed, only drugs recurrently identified across all DEG sets were considered. Finally, to obtain a single representative reversal score for each selected drug, the mean value across DEG sets was computed.

#### Combined score

A composite score was generated for drug prioritisation by integrating results from the signature-matching and network-analysis approaches. First, all individual scores were normalised to a 0–1 range to ensure comparability. Subsequently, the normalised scores were combined according to three criteria: (1) if a drug was identified by both approaches, the combined score was calculated as the sum of its signature-matching and network-analysis scores, plus two; (2) if a drug was identified only through signature matching, the combined score was defined as its signature-matching score plus one; (3) if a drug was identified solely through network analysis, the combined score remained unchanged. This scoring system prioritised drugs supported by both approaches while also favouring those identified by signature matching, which captures disease-specific gene expression patterns.

### Supplementary results

#### Exploratory analysis and correction of batch effects and the endometrial timing effect

Of the 20,802 genes initially detected, 5,465 were removed due to low expression, leaving 15,367 genes for downstream analyses. Batch effects were then evaluated, revealing a sequencing-run effect, which was subsequently corrected (**Supplementary Figure S3**).

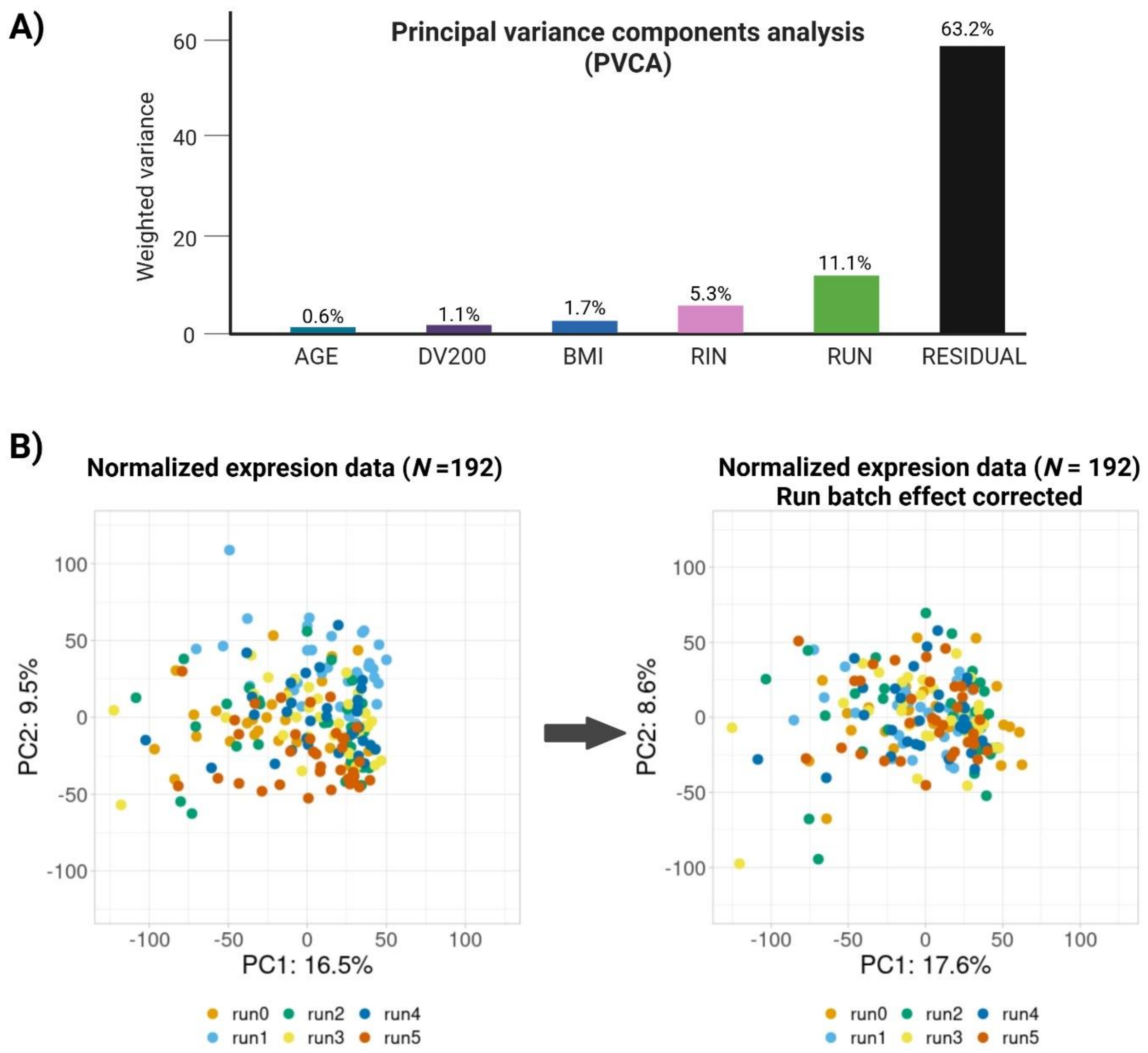


**Supplementary Figure S3**. **Analysis of potential batch effects. (A)** Weighted variance (%) associated with each potential batch effect. 'Residual' indicates the percentage of variance not explained by the evaluated variables. **(B)** Principal component analysis (PCA) of normalised gene-expression data before (left) and after (right) correction of batch effects associated with sequencing run. Abbreviations: BMI, body mass index; PC1, principal component 1; PC2, principal component 2.

Moreover, to remove the endometrial gene-expression variability caused by cyclic tissue changes (the timing effect), the TED model was applied. This identified 102 samples as pre-receptive (PR), 55 samples as receptive 1 (R1), 31 as receptive 2 (R2), and 4 as post-receptive (PS; **Supplementary Figure S4A**). The weighted variance associated with endometrial timing in principal variance components analysis (PVCA) was 12.4% (**Supplementary Figure S4B**) and therefore required correction (**Supplementary Figure S4C**).

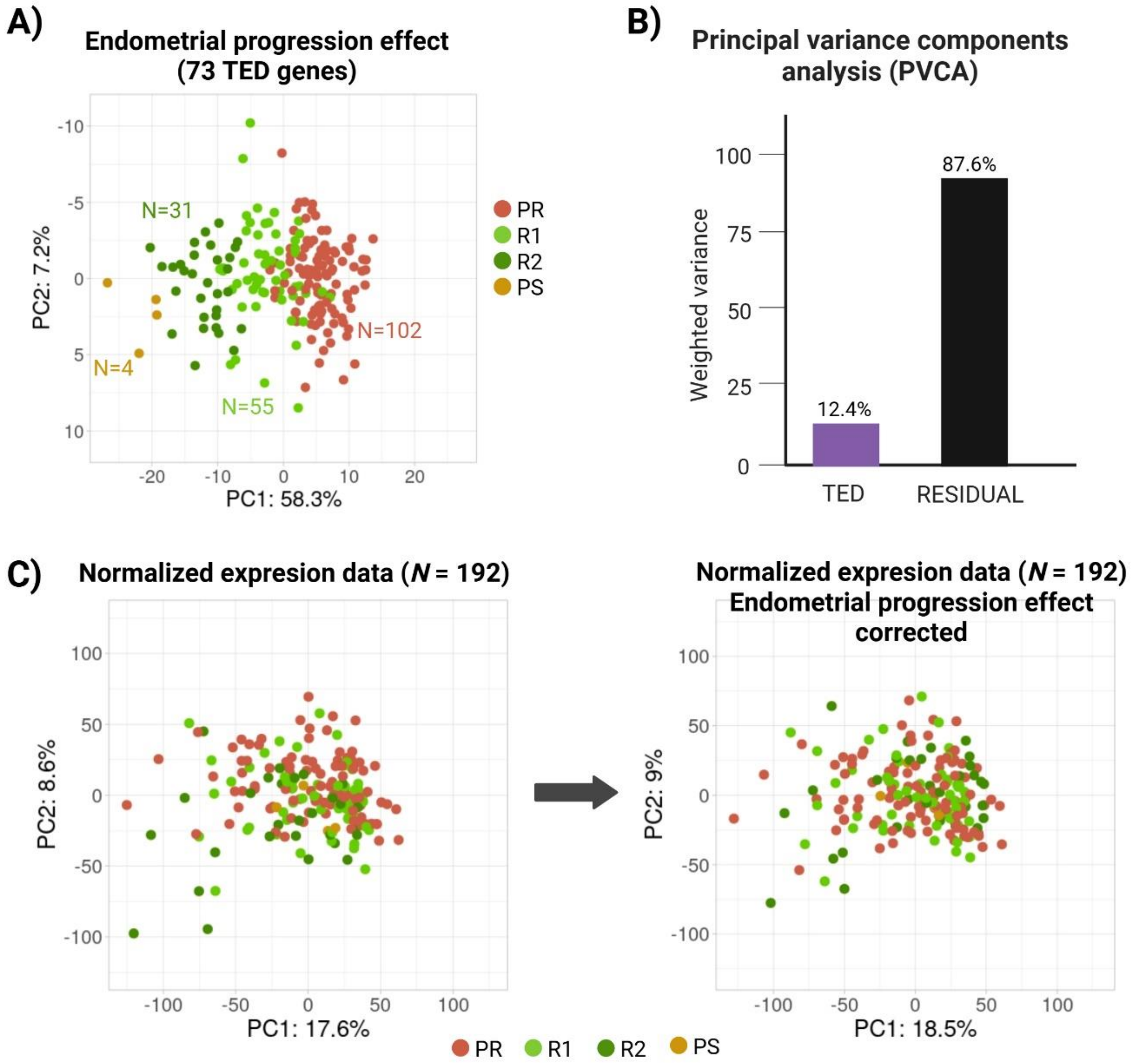


**Supplementary Figure S4**. **Evaluation of the batch effect associated with cyclic variation of the endometrium.** **(A)** Principal component analysis (PCA) plot showing the variance explained by 73 genes from the transcriptomic endometrial dating (TED) signature. **(B)** Principal variance component analysis (PVCA) showing the weighted variance (%) associated with the endometrial timing effect. 'Residual' indicates the percentage of variance not explained by the evaluated variables. **(C)** PCA plots showing correction of the endometrial timing effect: left, PCA of all the genes before correction; right, PCA after correction.

### Clinical results of the first stratification step

After applying  the semi-supervised learning algorithm described in Supplementary Methods through four iterations, 161 (83.9%) patients were successfully classified. Specifically, 43 patients were categorised as having a pathological-like (PL) endometrium and 118 as having a fertile-like (FL) endometrium based on their transcriptomic profiles, while 31 (16.1%) remained unclassified. Significant differences were observed between the PL and FL groups in terms of the following reproductive outcomes: pregnancy rate (26.3% vs. 66.0%, $p = 1.99 \times 10^{-5}$), live birth rate (40.0% vs. 87.1%, $p = 0.0013$), clinical miscarriage rate (42.9% vs. 6.2%, $p = 0.0159$), and biochemical miscarriage rate (30.3% vs. 7.2%, $p = 0.0372$).

**Additional functional analysis of transcriptomic profiles**

The remaining comparisons between profiles (CMA vs. PA, BMA vs. PA, and CMA vs. BMA) also revealed significant biological processes. The comparison between CMA and PA identified 45 significant functions, showing upregulation of hormonal response, metabolism, and signal transduction, together with downregulation of cellular respiration (**Supplementary Figure S5A**). When BMA was compared to PA, upregulated hormonal response and downregulated cellular respiration were observed, along with alterations in inflammatory and the immune-related pathways, further supporting immune dysregulation as a key feature of the BMA profile (**Supplementary Figure S5B**). Finally, when poor-prognosis profiles were compared with each other (CMA vs. BMA), 58 enriched functions were detected, primarily linked to metabolic activity, with the CMA profile showing increased metabolism and impaired cellular respiration (**Supplementary Figure S5C**).

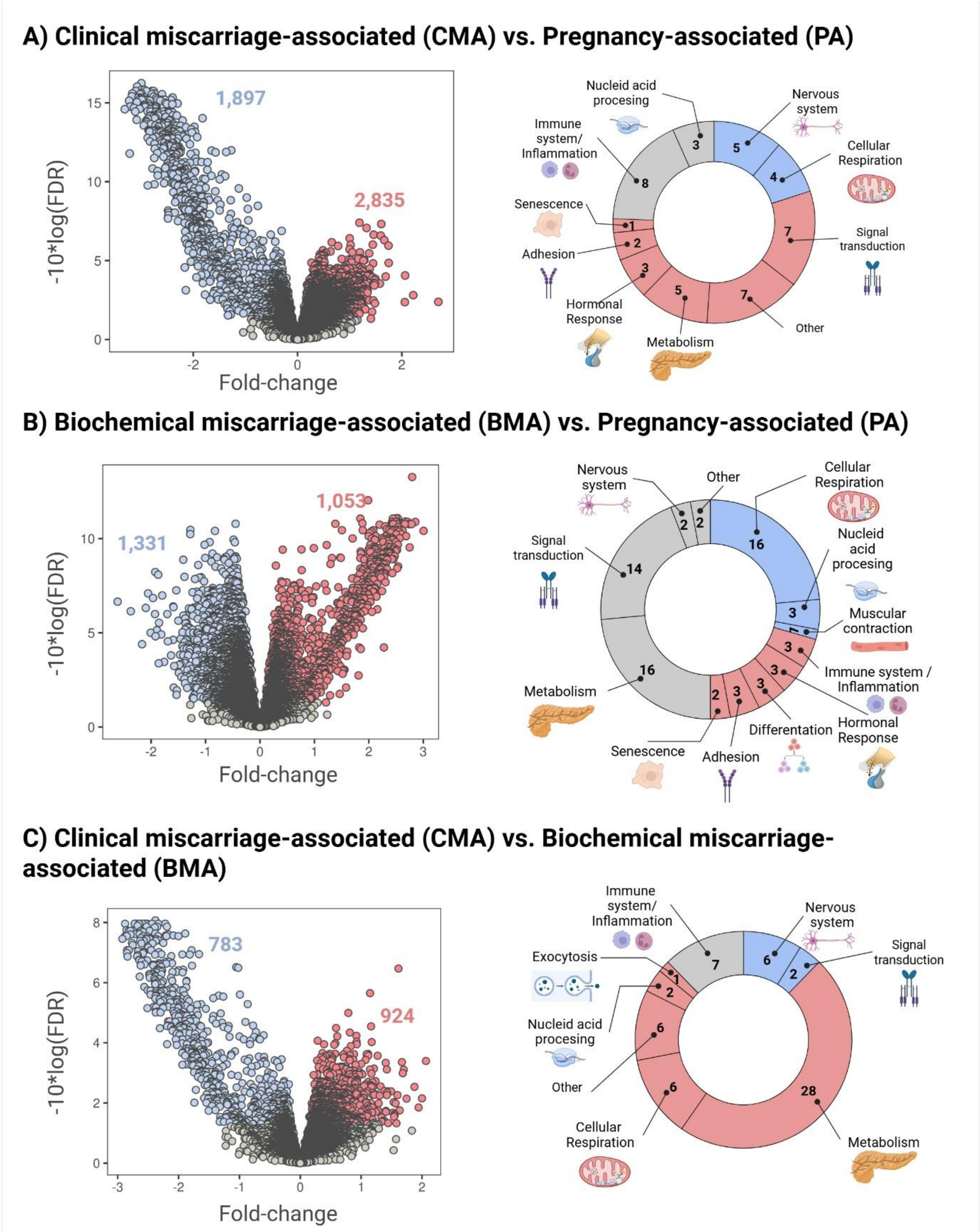


**Supplementary Figure S5: Transcriptomic and functional characterisation of molecular profiles of endometrial disruption. (A)** CMA vs. PA; **(B)** BMA vs. PA; **(C)** CMA vs. BMA. Volcano plots (left panels) represent the results of the differential expression analysis for each comparison. Colours indicate whether DEGs were significantly upregulated (red), significantly downregulated (blue), or non-significant (grey) when compared with the LBA profile. Doughnut plots (right panels) show the different biological functions identified through functional analysis for each molecular profile comparison. Colours indicate whether biological functions showed overall upregulation (red), overall downregulation (blue), or both (grey). All biological functions represented were statistically significant (FDR < 0.05). Abbreviations: BMA, biochemical miscarriage-associated; CMA, clinical miscarriage-associated; PA, pregnancy-associated; FDR: false discovery rate.

**Network and signature-matching analysis**

For the identification of effective and safe treatments for profiles associated with poor-prognosis (CMA and BMA), we focused on the DEGs obtained from their comparison with the gold-standard profile representing the healthiest endometrium, the live-birth-associated (LBA) profile. The CMA network comprised 151 genes, whereas the BMA network included 2,867 genes, reflecting the greater transcriptomic differences observed in the differential expression analysis. Both networks showed a scale-free distribution ($R^2$ = 0.85 for CMA and $R^2$ = 0.91 for BMA), supporting their biological network behavior.

Subsequently, these networks were used to identify all approved drugs with at least one target gene within the CMA or BMA networks. To ensure that the identified drugs were not selected by chance and to assess their potential therapeutic efficacy, a proximity analysis was applied, evaluating the closeness of each drug to the disease-related genes. After applying this analysis, only four drugs for CMA and 89 for BMA met the significance criteria and were selected for further prioritisation.

Regarding the signature matching approach, to avoid selecting an arbitrary number of genes as input for the algorithm, distinct sets of DEGs were generated according to the distributions of adjusted *p*-values and fold-change values. A total of 28 DEG sets were created for the CMA profile and 45 for the BMA one. These DEG sets were then used to identify potential drugs through the signature-matching approach. As a result, 480 and 346 significant drugs were identified when considering all DEG sets for CMA and BMA, respectively. Subsequently, only drugs detected in more than 7 DEG sets for CMA or 10 for BMA were retained for further analyses. These thresholds were determined according to data distribution (third quantile). Consequently, 55 significant drugs for CMA and 40 for BMA were selected for downstream analyses.

**Drug-dose selection**

To validate the mechanisms of action of selected treatments in endometrial tissue, a non-cytotoxic and physiologically relevant concentration was determined for each drug. Concentration ranges between 1 μM and 300 μM were tested in all cases, except terfenadine, which was assessed at lower concentrations (0.01 μM–1 μM). All drugs maintained a mean cell viability above 80% at specific concentrations (**Supplementary Figure S6**). Supplementary Table S4 lists the final concentrations selected for each drug.

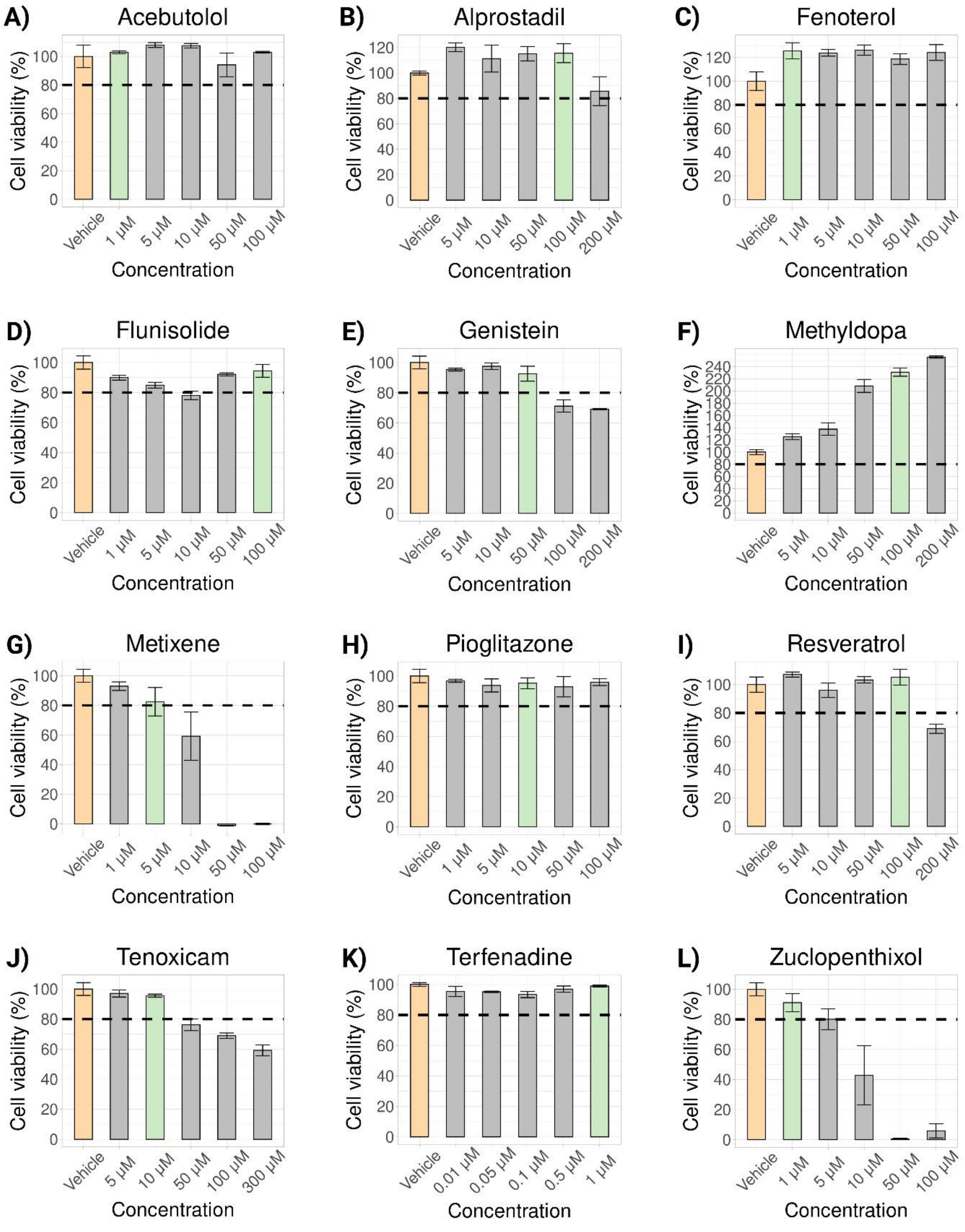


**Supplementary Figure S6. Cell viability assay for selected drugs.** Different concentrations of each drug were used to assess cell viability and determine non-cytotoxic doses. The *x*-axis represents drug concentration and the *y*-axis the percentage of cell viability. Dashed lines indicate the 80% viability threshold used for dose selection. The concentration is shown in light green and the vehicle in light orange. Error bars represent the standard error of the mean (SEM).

### Supplementary Table S4. Selected concentrations for in vitro validation.

| Drug | Concentration | Profile |
|---|---|---|
| Zuclopenthixol | 1 µM | CMA/BMA |
| Metixene | 5 µM | CMA/BMA |
| Genistein | 50 µM | CMA/BMA |
| Terfenadine | 1 µM | CMA/BMA |
| Methyldopa | 100 µM | CMA/BMA |
| Pioglitazone | 10 µM | CMA/BMA |
| Resveratrol | 100 µM | CMA/BMA |
| Flunisolide | 100 µM | BMA |
| Alprostadil | 100 µM | BMA |
| Tenoxicam | 10 µM | BMA |
| Acebutolol | 1 µM | BMA |
| Fenoterol | 1 µM | BMA |

The table lists the concentrations selected for testing each drug. The ‘profile’ column indicates whether the mechanism of action of each drug was observed in CMA, BMA, or both profiles. Abbreviations: CMA, clinical miscarriage-associated; BMA, biochemical miscarriage-associated.
